# Audiovestibular adverse events following COVID-19 vaccinations

**DOI:** 10.1101/2023.11.13.23298435

**Authors:** Aishwarya N Shetty, Hannah J Morgan, Linny K Phuong, John Mallard, Diana Vlasenko, Christopher Pearce, Nigel W Crawford, Jim P Buttery, Hazel J Clothier

## Abstract

**Importance:** Evidence regarding audiovestibular adverse events post COVID-19 vaccination to date has been inconclusive regarding a potential etiological association. This study used a multi-data source approach to assess incidence of these events following COVID-19 vaccination.

**Objective:** To determine if there was an increase in audiovestibular adverse events following COVID-19 vaccination in South-eastern Australia during January 2021 – March 2023.

**Design:** Retrospective observational analysis of spontaneous reports of audiovestibular events to a statewide vaccine safety surveillance service, SAEFVIC, as well as accompanying self-controlled case series (SCCS) analysis using general practice data collected via the POpulation Level Analysis and Reporting (POLAR) tool with permission from Primary Health Networks (PHNs) as the de-identified dataset owners in Victoria and New South Wales.

**Setting:** Victoria and New South Wales (NSW), Australia.

**Participants:** Victorians who spontaneously reported an audiovestibular-related symptom or diagnosis to SAEFVIC, and people in Victoria and NSW who presented to a POLAR GP registered practice with a new audiovestibular diagnosis.

**Exposures:** COVID-19 vaccination with adenovirus vector, mRNA or protein-subunit vaccine.

**Outcomes and Measures:** In SAEFVIC, audiovestibular events of interest were ascertained through searching key words in the vaccine safety database. Reporting rates were calculated and compared per 100,000 COVID-19 vaccine doses administered and recorded in the Australian Immunisation Register (AIR). Audiovestibular presentations of interest were isolated from the general practice dataset aggregated by POLAR, by searching for relevant SNOMED CT codes. Similarly, relative incidence (RI) was calculated for all COVID-19 vaccine types.

**Results:** This study demonstrates an increase in general practice presentations of vertigo following mRNA vaccines (RI= 1.40 *P* <.001), and tinnitus following both the adenovirus vector and mRNA vaccines (RI= 2.25, *P* <.001 and 1.53, *P* <.001 respectively). There was no increase in hearing loss following any COVID-19 vaccinations.

**Conclusions and Relevance:** This is the first study that demonstrates an increase in audiovestibular presentations following COVID-19 vaccination, in particular, vertigo and tinnitus. Healthcare providers and vaccinees should be alert to potential audiovestibular complaints after COVID-19 vaccination. Our analysis highlights the importance of using large real-world datasets to gather reliable evidence for public health decision making.

**KEY POINTS:** ***Question:*** Is there an increase in audiovestibular adverse events after COVID-19 vaccination (adenovirus vector [AstraZeneca’s Vaxzervria® ChadOx1-S], mRNA [Pfizer-BioNTech’s Comirnaty® BNT162b2 and Moderna’s Spikevax®] or protein-subunit [Novavax’s Nuvaxovid®])?

***Findings:*** This Australian study using spontaneous surveillance reports and large-scale general practice data, found an increase in incidence related to vertigo following mRNA vaccines (Relative Incidence = 1.40, *P* <.001), and tinnitus following both adenovirus vector and mRNA vaccines (Relative Incidence = 2.25, *P* <.001 and 1.53,*P* <.001 respectively). No increase in hearing loss following vaccination was observed.

***Meaning:*** Healthcare providers and vaccinees should be alert to potential audiovestibular complaints following COVID-19 vaccination.

## BACKGROUND

Following the implementation of COVID 19 vaccines commencing in December 2020^1^, concerns have been raised globally about a possible association between COVID-19 vaccinations and audiovestibular conditions ^2^. Australia’s COVID-19 vaccination rollout commenced on 22 February 2021^3^. The vaccine rollout began with mRNA Comirnaty BNT162b2 (Pfizer-BioNTech), followed by Adenovirus vector vaccine Vaxzevria ChadOx1-S (AstraZeneca), then mRNA Spikevax (Moderna) and protein-based vaccine Nuvaxovid (Novavax)^1, 4^. More recently, bivalent variations of COVID-19 mRNA vaccines have also been introduced as part of a booster program^1^.

Audiovestibular events relate to a broad spectrum of signs and symptoms including hearing loss, tinnitus, and vertigo. *Hearing loss* is characterized by the inability to hear sound in one or both ears^5^, *vertigo* is characterized by a sudden internal spinning sensation^6^, and *tinnitus* is characterized by perception of sound without an external auditory stimulus^7^. Although not life threatening, these adverse events may significantly impact the quality of life of those affected, cause sociopsychological distress and can severely impact vaccine confidence in the community ^8^.

Clinical trials did not show increased audiovestibular events in the vaccine arm^9^. Furthermore, a majority of published literature to date includes case studies, case series and spontaneous report studies, which are prone to bias and do not provide sufficient comparative evidence to confidently support or refute a causal relationship with prior vaccination^2, 10^. This population-wide study uses a multiple data source approach to determine if the incidence of audiovestibular conditions are increased following COVID-19 vaccinations.

## METHODS

This analysis included two methodologies. First, an epidemiological review of audiovestibular events following COVID-19 vaccination reported to a state-level vaccine safety spontaneous reporting platform (SAEFVIC). Second, a self-controlled case series (SCCS) analysis conducted on a large general practice (GP) dataset to explore if there was an increase in audiovestibular presentations in the 42-day risk window post COVID-19 vaccination compared with other periods.

### Surveillance of Adverse Events Following Immunisation in the Community (SAEFVIC) report analysis

SAEFVIC is the central spontaneous reporting service for adverse events following immunization (AEFI) in the Australian state of Victoria (population 6.5 million). SAEFVIC is responsible for epidemiological data analysis and referral of vaccinees with significant AEFIs to specialist vaccination clinics where required. Surveillance comprises enhanced passive and active surveillance systems integrated with clinical services^11^. For COVID-19, spontaneous reports were received from healthcare professionals and/or vaccinees, or via AusVaxSafety solicited state government text message surveys on day 8 and day 42 post-vaccination^12^.

The analysis used a consensus list of keywords describing audiovestibular events based upon Brighton Collaboration Criteria and applicable ICD-10-AM (International Statistical Classification of Diseases – Tenth Revision – Australian Modification) diagnoses^13^. The identified keywords were grouped to three broad categories – hearing loss, vertigo, and tinnitus (eTable 1). Reports matching these keywords submitted to SAEFVIC between 22 February 2021 and 28 March 2023 were identified and then filtered to only include adults aged between 18-65 years of age who had an audiovestibular symptom onset within 42 days of vaccination. Timing of symptom onset was determined by the cases self-reported symptom onset date within the report, or where this was missing, the report submission date.

Other information contained within reports and used for the analysis included case demographics including sex and age; vaccination details including date of vaccination, COVID-19 vaccine brand and dose number; and clinical characteristics including symptomology, and time to onset of symptoms.

AEFI count and reporting rates were calculated per 100,000 doses administered as recorded in the Australian Immunisation Register (AIR)^14^ using Microsoft PowerBI (version 2.91.701.0) and 95% Poisson confidence intervals calculated and p values calculated using RStudio (version 4.2.3).

### Outcome Health’s POpulation Level Analysis and Reporting (POLAR) platform

The POLAR platform collects and processes general practice data on behalf of Primary Heath Networks (PHNs). The de-identified dataset, owned by the source PHN respectively, contains over 12 million case records from PHNs in Victoria and New South Wales^15^ namely: Central and Eastern Sydney PHN, Eastern Melbourne PHN, Gippsland PHN, South Eastern Melbourne PHN, and South Western Sydney PHN.

The POLAR dataset was analysed using a SCCS epidemiological study design. In an SCCS design, each case acts as their own control in their pre- and post-vaccination window. This limits the need for recruiting controls and time invariant confounding factors are controlled for^16^. The SCCS analysis is particularly useful for investigating the association between transient exposures like vaccines and adverse events.

We applied the SCCS methodology to determine if there was an increase in relative incidence of audiovestibular presentations in the period immediately post COVID-19 vaccination (risk window) as compared to baseline periods. The study period included people aged 20-65 years with an event presentation from 1 January 2021 to 28 March 2023.The risk window was defined as 1-42 days following vaccination with any COVID-19 vaccine (excluding day 0, the day of vaccine administration, according to the biological plausibility of a vaccine-related audiovestibular diagnosis. All other time periods other than the risk window and within the study duration constituted the baseline period (control period) (Figure 1). We only included the first time a person presented with an audiovestibular condition in the observation period and all subsequent audiovestibular visits were excluded. The SCCS analysis outcome SNOMED CT codes were grouped into three broad categories – hearing loss, tinnitus, and vertigo.

**Figure 1:**
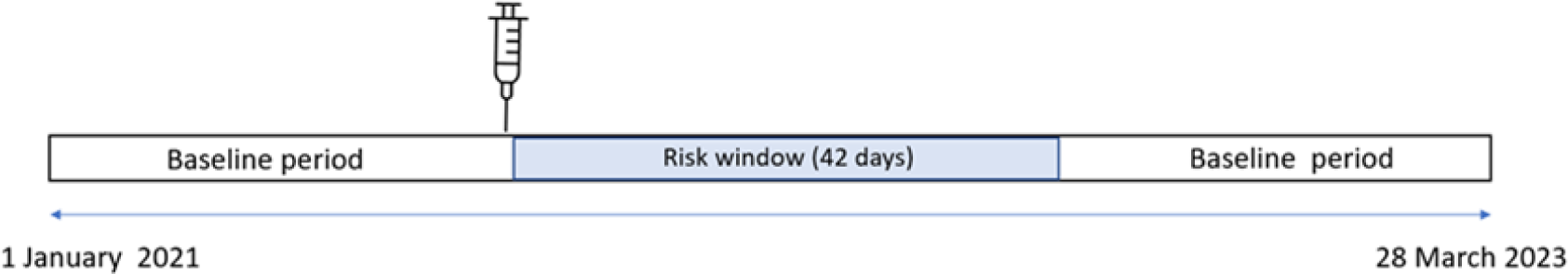
Observation period and risk window applied in the SCCS methodology.

Descriptive and statistical analyses were conducted in the POLAR secure data environment using Q ik Sense analytics platform (Qlik Technologies, PA) and the *SCCS* Package in RStudio (version 4.2.3)^17,18, 19^.

### Sub analysis

All analyses were conducted collectively, as well as by vaccine type and dose number. Analyses were conducted for all individual conditions and as three audiovestibular groups: hearing loss, vertigo, and tinnitus.

### Exclusions

Those symptoms and diagnoses in which pathogenesis is attributable to other pathology, such as an intracranial brain lesion, neck pathology or ischaemic stroke, were excluded. Such terms included endolymphatic hydrops and Meniere’s disease which were excluded from both datasets. The following terms were excluded from the general practice dataset (and were not present in the SAEFVIC database): cervical vertigo; malignant positional vertigo; vertebrobasilar ischaemic vertigo and tinnitus of vascular origin.

Due to insufficient case records, reports for the following vaccines were excluded from the analysis: Nuvaxovid® (SAEFVIC=3 reports, GP data=41 risk-window presentations), mRNA based-Spikevax® monovalent (SAEFVIC=29, GP data=not available), and bivalent mRNA vaccines (SAEFVIC=1, GP data=not available).

### Ethics

Access to SAEFVIC data conforms to the function as public health vaccine safety surveillance approved by the Royal Children’s Hospital Health Research Ethics Committee as a registered database # 36219. The POLAR platform has ethics approval for collection, transfer, and storage of general practice data (RACGP NREEC Protocol ID: 17-008) Ethics approval for use of general practice data collected and processed by POLAR for this project was provided by Monash Human Research Ethics Committee [RES-18-0000-232A].

#### Data availability

The SAEFVIC data used to generate counts and rates is collected from vaccinees and patients with adverse events as part of routine public health surveillance practices. The SAEFVIC dataset contains sensitive identifiable information. As such, the dataset access is not publicly available and is restricted to select SAEFVIC employees in a secure environment. The data that support the SCCS findings of this study are hosted by POLAR, but restrictions apply to the availability of these data, which were used under license for the current study, and so are not publicly available. All the data used for the SCCS analysis is completely deidentified.

## RESULTS

### Data summary

The study reviewed 45,350 reports and 4,940,000 presentations recorded in the SAEFVIC and the general practice datasets respectively between 1 January 2021-28 March 2023. On application of the eligibility criteria, the most frequently reported audiovestibular condition was vertigo (SAEFVIC: 415, GP data: 13,924) followed by tinnitus (SAEFVIC: 226, GP data: =4,000) and hearing loss (SAEFVIC: 76, GP data: 3,214) (Figure 2) (eTable 1).

**Figure 2:**
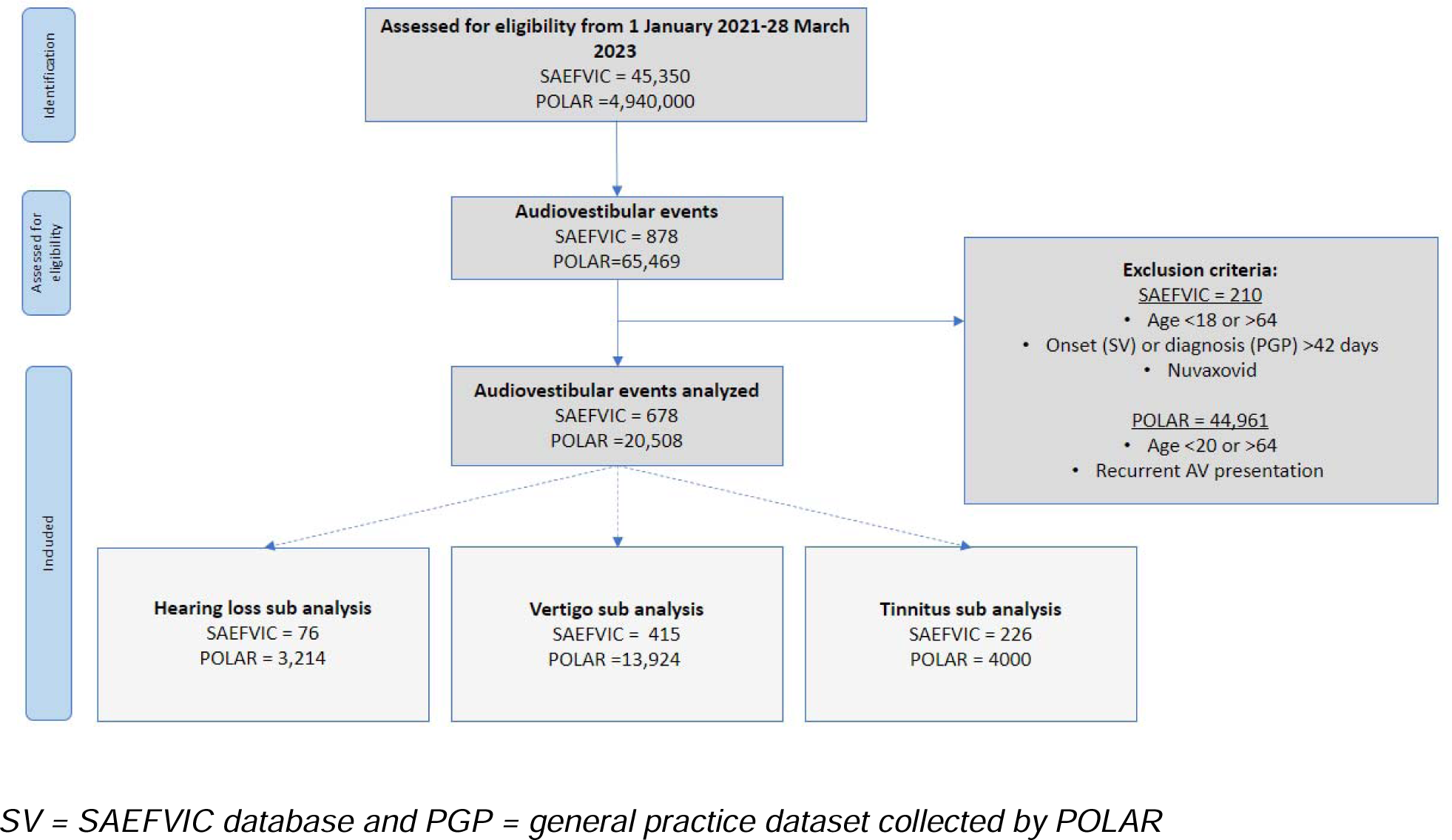
Identification of reports for inclusion and analysis.

### All audiovestibular conditions

678 spontaneous reports of audiovestibular conditions were received following adenovirus vector-based and mRNA-based vaccines in adults between 18–64 years with an onset ≤42 days. The reporting rate was higher for adenovirus vector-based vaccines compared to mRNA vaccines (rate ratio: 1.94, 95%CI 1.65, 2.23) (Table 1). Rates were higher following dose 1 compared to dose 2 for the adenovirus vector-based vaccine (rate ratio: 4.1, 95%CI 2.89, 5.87) but no difference in incidence rates by dose received were observed for mRNA vaccines (dose 1 to dose 2 rate ratio: 1.02, 95%CI 0.84, 1.25) (Table 2).

**Table 1:** Summary reporting rates and relative incidences of audiovestibular conditions by COVID-19 vaccine type.

|  | SAEFVIC: Rate per 100,000 (95% CI<sup>a</sup>) |  |  |  | GP data via POLAR: Relative incidence (95%CI<sup>a</sup>, *P*-value) |  |  |  |
| --- | --- | --- | --- | --- | --- | --- | --- | --- |
| Condition | Total | Adenovirus vector (Vaxzevria) | mRNA (Comirnaty and Spikevax) | Rate ratio Adenovirus vector/mRNA | Total | Adenovirus vector (Vaxzevria) | mRNA (Comirnaty and Spikevax) | Summary finding(s) |
| All audiovestibular | 5.8 (5.4, 2.3) | 9.7 (8.3, 11.2) | 5.0 (4.6, 5.5) | 1.94 (1.65, 2.23) (*P* <.001) | 1.39 (1.28, 1.51, *P* <.001) | 1.36 (1.08, 1.71, *P* <.01) | 1.38 (1.26, 1.51, *P* <.001) | Higher reporting rate for adenovirus vector vaccine and SCCS signal post both vaccine types |
| Hearing loss | 0.6 (0.5, 0.8) | 0.9 (0.5, 1.4) | 0.6 (0.4, 0.7) | 1.5 (0.9,2.1) (*P* =.2) | 1.11 (0.88, 1.4, *P* =.35) | 1.08 (0.61, 0.93, *P* =.776) | 1.13 (0.88, 1.45, *P* =.316) | No signal |
| Vertigo | 3.6 (3.2,3.9) | 5.9 (4.9, 7.1) | 3.1 (2.8, 3.5) | 1.90 (1.56,2.24) (*P* <.001) | 1.41 (1.27, 1.55, *P* <.001) | 1.23 (0.92,1.65, *P* =.15) | 1.40 (1.26, 1.56, *P* <.001) | Higher reporting rate for adenovirus vector vaccine and SCCS signal post mRNA vaccines |
| Tinnitus | 1.9 (1.7, 2.2) | 3.2 (2.5, 4.1) | 1.7 (1.4, 2.0) | 1.88 (1.32, 2.44) (*P* <.001) | 1.60 (1.3, 1.93, *P* <.001) | 2.25(1.45, 3.50, *P* <.001) | 1.53 (1.25, 1.87, *P* <.001) | Higher reporting rate for Adenovirus vectored vaccine and SCCS signal post both vaccine brands |

95%CI<sup>a</sup> = 95% Confidence Interval

**Table 2:**
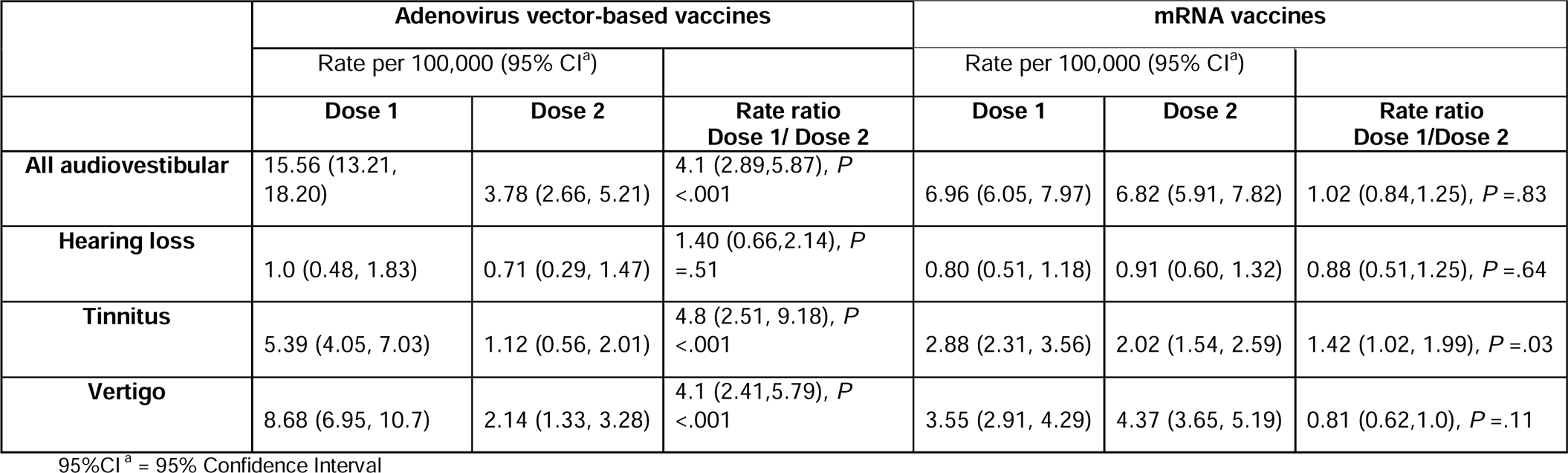
Summary rates and 95% confidence intervals of audiovestibular conditions by COVID-19 vaccine dose number and vaccine type.

SCCS analysis demonstrated increased risk of audiovestibular conditions in the 42 days following adenovirus vector (relative incidence: 1.36, 95%CI 1.08, 1.71) and mRNA vaccines (relative incidence: 1.38, 95%CI 1.26, 1.51) (Table 1).

In the SAEFVIC database, the reporting rate for females was greater than males (Rate ratio: 3.5, *P*<.001) and the median age of audiovestibular reports was 41 years (interquartile range: 19) (eTable 2). In the general practice data aggregated by POLAR, there was no differences in audiovestibular presentations between sexes (relative incidence ratio: 1.08, 95%CI 0.92, 1.24 *P* =.2) and no age-disaggregated data were available.

### Sub-analysis groups

#### Hearing loss

Seventy-six spontaneous reports of hearing loss were received, of which 32% (24/76) were of sudden onset (within 3 days). No differences in reporting rates were identified by vaccine type (rate ratio of adenovirus vector-based vaccines compared to mRNA vaccines: 1.5, 95%CI 0.9, 2.1). No statistically significant differences in reporting rate between doses were observed (adenovirus vector dose 1 compared to dose 2: rate ratio 1.40, 95%CI 0.66, 2.14; mRNA dose 1 compared to dose 2 rate ratio: 0.88, 95%CI 0.51, 1.25) (Table 2).

SCCS analysis also did not find any increased risk of hearing loss in the 42 days following either vaccine type (adenovirus vector Relative incidence: 1.08, 95%CI 0.61, 0.93; mRNA relative incidence: 1.13, 95%CI 0.88, 1.45). (Table 1)

#### Vertigo

415 spontaneous reports of vertigo were submitted to SAEFVIC. Reporting rates of vertigo were higher following adenovirus vector vaccine compared to mRNA vaccines (rate ratio: 1.9, 95%CI 1.56,2.24). Rates were higher following dose 1 compared to dose 2 for the adenovirus vector vaccine (rate ratio: 4.1, 95%CI 2.41,5.79) but no differences in reporting rate between doses were observed for mRNA vaccines (dose 1 compared to dose 2 rate ratio: 0.81, 95%CI 0.62,1.0) (Table 2).

SCCS analysis demonstrated a statistically increased risk of vertigo in the 42 days following mRNA vaccines (relative incidence: 1.40, 95%CI 1.26, 1.56) compared to baseline periods (Table 1).

#### Tinnitus

226 spontaneous reports of tinnitus were submitted to SAEFVIC. Reporting rates of tinnitus were higher following adenovirus vector vaccine compared to mRNA vaccines (rate ratio: 1.88, 95%CI 1.32, 2.44). Rates were higher following dose 1 compared to dose 2 for both the adenovirus vector vaccine (rate ratio: 4.8, 95%CI 2.51, 9.18) and mRNA vaccines (rate ratio: 1.42, 95% CI 1.02, 1.99) (Table 2).

SCCS analysis demonstrated an increased risk of tinnitus in the 42 days following both adenovirus vector vaccine (relative incidence: 2.25, 95%CI 1.45, 3.50) and mRNA vaccines (relative incidence: 1.53, 95%CI 1.25, 1.87) (Table 1) compared to baseline periods.

## DISCUSSION

We investigated the proposed association between COVID-19 vaccination and audiovestibular conditions by reviewing statewide data gathered between January 2021 and March 2023. Reporting patterns were analysed using spontaneous reports received by SAEFVIC, and the analysis was further strengthened by conducting a SCCS analysis using general practice data collected via the POLAR platform. Our study found an increased relative incidence of vertigo in the 42 days following mRNA vaccines, and an increased relative incidence of tinnitus in the 42 days following both adenovirus vector and mRNA vaccines. We are the first to confirm this increased relative incidence of tinnitus and vertigo post COVID-19 vaccines ^20, 21^. Importantly, no increased relative incidence in hearing loss was observed in the 42 days following any COVID-19 vaccine, supporting results described previously ^22, 23, 24^,

Although the pathophysiology of audiovestibular events following vaccination is unclear, proposed mechanisms include an immune mediated injury due to exaggerated cytokine responses leading to vasculitic events, antibody cross-reactivity and molecular mimicry^2^. Another suggested mechanism is that mRNA vaccines can cause reactivation of previous latent viruses resulting in sudden hearing loss^25^. Of significance, COVID-19 infection itself has also been linked to audiovestibular events and there were over 11,000,000 cases of COVID-19 infection during our study period^26^. COVID-19 prior infection status and symptom onset dates are unknown for the cases included in this study, but it is highly likely that many of our cases had experienced COVID-19 infection at a similar time to their vaccination and audiovestibular events. Therefore, COVID-19 infection is an important potential confounder of the association between COVID-19 vaccination and audiovestibular events ^21, 27^.

Our analysis supports the opinion that there is no increased incidence of hearing loss following COVID-19 vaccines. Consistent with our findings, the US Vaccine Adverse Event Reporting System (VAERS) data, and studies conducted on the Finnish and Danish health care registry, found no association between sudden sensorineural hearing loss (SSNHL) and COVID-19 vaccination^22, 23, 24^.

Notably, our analysis demonstrated conflicting results across the two datasets under investigation. The SAEFVIC spontaneous reports show a greater reporting rate of vertigo reports following adenovirus vector compared to mRNA vaccines, but the SCCS analysis conducted on the primary care dataset indicated a signal post-mRNA vaccine, and none following adenovirus vector vaccines. A possible explanation for this could be that adenovirus vectored vaccines in Australia were predominantly administered in an older cohort at greater risk of vertigo, which may lead to reporting bias in SAEFVIC for adenovirus vector vaccines. As only new diagnoses were included for a patient in the primary care SCCS analysis, exacerbations of vertigo may have been reported spontaneously, but excluded from the SCCS population. In contrast, a small retrospective case series (n=33) of COVID-19 infection naïve cases demonstrated no statistically significant association of acute vertigo to mRNA-based COVID-19 vaccination^21^.

Our analysis demonstrated an increased incidence of tinnitus for all vaccine types, with a greater incidence following adenovirus vector vaccines. Of interest, in those individuals with pre-existing tinnitus, studies during the COVID-19 pandemic demonstrated that environmental stress may contribute to tinnitus^27^. Contrary to our findings, a study conducted on the Federated Health Data Network in the US found the rate of newly diagnosed tinnitus three weeks after the first dose of COVID-19 vaccine was very low^20^. Similarly, a retrospective chart review found no definite correlation between tinnitus occurring within four weeks of COVID-19 vaccination^28^. However, most of these studies rely on case self-reports and are prone to recall bias. Two papers describing four case reports of tinnitus following COVID-19 vaccination had onset times of five hours after adenovirus vector vaccine (Vaxzevria) and seven hours to six days following mRNA vaccine (Comirnaty)^29, 30^. Three of the four cases were in persons aged between 30-63 years of age, all were male and two had a history of autoimmune conditions.

Our spontaneous reporting platform demonstrated four-times higher reporting rate of audiovestibular adverse events following dose 1 adenovirus vector vaccines compared to dose 2. This was not unexpected as there is established increased general reactogenicity to dose 1 of adenoviral vector vaccines^31^. Our SCCS analysis was unable to assess this finding as dose sequence number is not completely available in the general practice data extraction, therefore precluding dose stratification analyses. Further studies will be needed to examine the risk of subsequent doses, including boosters, on audiovestibular conditions as well as flares for those with existing conditions.

Literature suggests anatomic variances in the inner ear do exist between sexes and our spontaneous reports detected a higher reporting rate in females^32^. A hypothesis for an increased prevalence of vertigo in females can be attributed to hormonal differences, gene polymorphism and myelination of vestibular nerve. ^32^ For tinnitus it has been found that females had higher tinnitus annoyance, which could possibly influence reporting behaviors. However, it is important to note, episodic dizziness (due to conditions including tension syndrome, menopause or even caloric restriction and emotional distress) can often be reported as vertigo when it comes to self-reporting^32^. Our SCCS analysis, which uses clinician diagnoses and controls for time invariant confounding factors, demonstrated negligible difference between sexes in audiovestibular diagnosis.

Uniquely, the strength of our study is its inclusion of SCCS methodology using healthcare seeking datasets, in addition to review of routine reporting datasets. This general practice dataset avoids the reporting biases associated with voluntary spontaneous AEFI reported conditions and is likely to be a more accurate representation of the true state of audiovestibular events in the community. In Australia, most patients consult a general practitioner first, as specialists require a medical referral. In addition, it is well known that individual spontaneous vaccine surveillance reports are not designed to assign causality due to the inability to adjust for confounders such as comorbidities or previous audiovestibular history, and a lack of complete records^33^. Spontaneous reports may also lack full clinical description of the reaction event, making assignment of and classifications within case definitions challenging. Thus, the multi-data source approach described in this study allows for more rigorous evaluation to provide an evidence base to inform policy decisions.

A limitation of our SCCS dataset is that it will not capture all COVID-19 vaccinations administered as a person can have their vaccination at any healthcare setting and not just those sites registered on the POLAR platform. This means that we could be assigning audiovestibular events to the baseline period, when in fact they may have been in the 42 days following a vaccine (risk window), but this information was not known. Such instances would lead to an underestimate of the true relative incidence of audiovestibular events.

Similar to our analyses, previous studies compare background incidence rates of these events to rates during COVID-19 vaccination rollout periods. It is difficult to assign causality of these events to vaccination due to the multiple concurrent variables contributing to these events and the lack of complete records for each of these affected cases.

## CONCLUSION

Our analysis used a combination of spontaneous-reports and general practice consultations data to assess any association between COVID-19 vaccines and audiovestibular conditions. We found an increase in vertigo presentations following COVID-19 mRNA vaccines, an increase in tinnitus presentations following COVID-19 mRNA vaccines and adenovirus vector vaccines, but no increase in hearing loss presentations following any COVID-19 vaccine. The multi-dataset combination method approach allowed us to interpret the results from each of the datasets helping minimise the biases introduced by them. Healthcare providers should be aware of these potential adverse events to ensure that people experiencing them, as well as those considering COVID-19 vaccines in the future, are well counselled.

## Data Availability

The datasets analysed for the current study are not publicly available.

## Acknowledgements

We thank Mr. Adam McLeod and Karina Gardner of Outcome Health, Melbourne, Victoria, for their support with this research for assistance with mapping of SNOMED CT codes. They did not receive any compensation for these roles.

## Author Contributions

AS had access to POLAR data and HM to SAEFVIC data and take responsibility for the integrity of the data and the accuracy of the data analysis.

Concept and design: All authors

Acquisition, analysis, or interpretation of data: All authors.

Drafting of the manuscript: AS, HM, LP, HC

Critical revision of the manuscript for important intellectual content: All authors

Statistical analysis: AS, HM

Administrative, technical, or material support: AS, HM

Supervision: JB, HC

