## Supplemental Tables for "Audiovestibular adverse events following COVID-19 vaccinations"

**eTable 1: Count of presentations by SNOMED code (POLAR) and reports by keyword (SAEFVIC) for each audiovestibular category**

|  | **GP Data via POLAR** | | | | **SAEFVIC** | | |
| --- | --- | --- | --- | --- | --- | --- | --- |
| **Category** | **SNOMED Code** | **SNOMED Text** | **Included presentation count** | **Total presentation count** | **Adverse event code** | **Included report count** | **Total report count** |
| All audiovestibular |  |  | 20508 | **65,469** |  | **678 reports presenting with 717 AV conditions** | **878 reports presenting with 894 AV conditions** |
| Hearing loss | **Total** | | **3214** | **16,664** | **Total** | **76** | **98** |
|  | 15188001 | Hearing loss |  |  | Hearing loss | 65 | 85 |
|  | 60700002 | Sensorineural hearing loss |  |  | Sensorineural hearing loss | 6 | 7 |
|  | 194424005 | Sensorineural hearing loss, bilateral |  |  | Complete hearing loss | 0 | 1 |
|  | 77507001 | Mixed conductive & sensorineural hearing loss |  |  |  |  |  |
|  | 73371001 | Neural hearing loss |  |  |  |  |  |
|  | 95820000 | Bilateral hearing loss |  |  |  |  |  |
|  | 473423001 | Hearing loss of right ear |  |  |  |  |  |
|  | 473424007 | Hearing loss of left ear |  |  |  |  |  |
|  | 103276001 | Decreased hearing |  |  |  |  |  |
|  | 128540005 | Hearing disorder |  |  | Transient hearing loss | 4 | 4 |
|  | 162340000 | Hearing difficulty |  |  | Altered hearing | 1 | 1 |
|  | 300228004 | Hearing problem |  |  |  |  |  |
|  | 362989003 | Auditory dysfunction |  |  |  |  |  |
|  |  | Obscure auditory dysfunction |  |  |  |  |  |
|  | 162344009 | Bilateral deafness |  |  |  |  |  |
|  | 343087000 | Partial deafness |  |  |  |  |  |
|  | 473421004 | Deafness of right ear |  |  |  |  |  |
|  | 473422006 | Deafness of left ear |  |  |  |  |  |
|  | Items considered, but not included: Ménière's disease, Endolymphatic hydrops, Otosclerosis, Auditory processing disorder | | | | | |  |
| Vertigo | **Total** | | **13294** | **38,729** | **Total** | **415** | **528** |
|  | 399153001 | Vertigo |  |  | Vertigo | 364 | 425 |
|  | 111541001 | Benign paroxysmal positional vertigo |  |  | Benign Paroxysmal Positional Vertigo: | 10 | 11 |
|  | 50438001 | Peripheral vertigo |  |  |  |  |  |
|  | 103284002 | Positional vertigo |  |  |  |  |  |
|  | 103290003 | Paroxysmal vertigo |  |  |  |  |  |
|  | 103291004 | Intermittent vertigo |  |  |  |  |  |
|  | 103298005 | Severe vertigo |  |  |  |  |  |
|  | 232284007 | Migrainous vertigo |  |  | Vestibular migraine | 6 | 6 |
|  | 20425006 | Labyrinthine disorder |  |  |  |  |  |
|  | 23919004 | Labyrinthitis |  |  |  |  |  |
|  | 445053006 | Dysfunction of vestibular system |  |  | Vestibular disorder flare | 1 | 1 |
|  |  | Impairment of balance |  |  | Loss of balance | 18 | 28 |
|  |  | Poor balance |  |  |  |  |  |
|  | 298313002 | Problem with balance |  |  |  |  |  |
|  |  | Unable to balance |  |  |  |  |  |
|  |  |  |  |  | Vestibular disturbance | 3 | 4 |
|  |  |  |  |  | Vestibular syndrome | 1 | 1 |
|  |  |  |  |  | Persistent postural perceptual dizziness | 2 | 2 |
|  |  |  |  |  | Peripheral vestibulopathy | 1 | 1 |
|  |  |  |  |  | Vestibular neuritis | 19 | 20 |
|  | Items considered, but not included: Ataxia, Cerebellar vertigo, Cervical vertigo, Recurrent labyrinthitis, Epidemic vertigo, Malignant positional vertigo, Vertebrobasilar ischaemic vertigo, Menieres Disease | | | | | | |
| Tinnitus | **Total** | | **4000** | **10,076** | **Total** | **226** | **268** |
|  | 60862001 | Tinnitus |  |  | Tinnitus | 226 | 268 |
|  | 162349004 | Noises in ear |  |  |  |  |  |
|  | 162351000 | Buzzing in ear |  |  |  |  |  |
|  | 162352007 | Ringing in ear |  |  |  |  |  |
|  | Items considered, but not included: Tinnitus of vascular origin | | | | | | |

**eTable 2: Demographic characteristics of reported cases of audiovestibular AEFI following COVID-19 vaccination in SAEFVIC.**

|  | Adenovirus vector | | | mRNA | | | **Totals** | | |
| --- | --- | --- | --- | --- | --- | --- | --- | --- | --- |
|  | **AEFI Count** | **Dose number** | **Rate per 100,000 (95%CI)** | **AEFI Count** | **Dose number** | **Rate per 100,000 (95%CI)** | **AEFI Count** | **Dose number** | **Rate per 100,000 (95%CI)** |
| **Age group** | | | | | | | | | |
| **18-19** | 1 | 31,979 | 3.1 (0.1-17.4) | 2 | 348,041 | 0.6 (0.1-2.1) | **3** | **380,020** | **0.8(0.2-2.3)** |
| **20-29** | 12 | 251,114 | 4.8(2.5-8.3) | 72 | 2,098,432 | 3.4 (2.7-4.3) | **84** | **2,349,546** | **3.6(2.9-4.4)** |
| **30-39** | 25 | 258,366 | 9.7(6.3-14.3) | 107 | 2,496,390 | 4.3 (3.5-5.2) | **132** | **2,754,756** | **4.8(4.0-5.7)** |
| **40-49** | 28 | 127,345 | 22.0(14.6-31.8) | 189 | 2,330,167 | 8.1 (7.0-9.4) | **217** | **2,457,512** | **8.8(7.7-10.1)** |
| **50-59** | 85 | 739,233 | 11.5(9.2-14.2) | 97 | 1,721,607 | 5.6(4.6-6.9) | **182** | **2,460,840** | **7.4(6.4-8.6)** |
| **60-69** | 43 | 592,100 | 7.3(5.3-9.8) | 17 | 625,872 | 2.7(1.6-4.3) | **60** | **1,217,972** | **4.9(3.8-6.3)** |
| **Sex** | | | | | | | | | |
| **Female** | 126 | 953,850 | 13.2(11.0-15.7) | 337 | 5,007,741 | 6.7(6.0-7.5) | **463** | **5,961,591** | **7.8(7.1-8.5)** |
| **Male** | 68 | 1,041,802 | 6.5(5.1-8.3) | 147 | 4,601,695 | 3.2(2.7-3.8) | **215** | **5,643,497** | **3.8(3.3-4.4)** |
| **Total** | **194** | **2,000,137** | **9.7(8.4-11.2)** | **484** | **9,620,509** | **5.0(4.6-5.5)** | **678** | **11,620,646** | **5.8(5.4-6.3)** |
